# Protocol for a scoping review of theorised pathways between home energy decarbonisation and health and health inequalities

**DOI:** 10.1101/2024.08.23.24312045

**Authors:** Vicki Ponce Hardy, Amy Stevenson, Gerry McCartney, Alison Heppenstall, Petra Meier

**Affiliations:** MRC/CSO Social and Public Health Sciences Unit, School of Health and Wellbeing, University of Glasgow, Glasgow, UK; School of Social and Political Sciences, University of Glasgow, Glasgow, UK; Urban Analytics, School of Social and Political Sciences, University of Glasgow, Glasgow, UK

**Keywords:** health, health inequalities, energy, energy efficiency, pathways, decarbonisation

## Abstract

**Introduction:** Access to adequate energy in the home is necessary for a healthy and well life, however current energy use, particularly in high-income countries, is unsustainable. Decarbonisation of home energy can benefit climate mitigation and health but there is the potential to create new, or compound existing, inequalities in health if not implemented equitably. Mapping the theoretical causal pathways between home decarbonisation and health will contribute to further understanding of these mechanisms.

**Aims:** Firstly, to identify theoretical pathways between decarbonisation of home energy and health and health inequalities in high-income countries, and secondly, to synthesise these into a putative causal evidence map.

**Inclusion criteria:** All populations in high-income countries are included, as defined by the World Bank in 2023/24. Included concepts are decarbonisation of home energy, and health and health inequalities. Context for this review comprises of the inclusion of a clear theory linking the concepts. All study designs are included.

**Methods:** This protocol is for a review of theories rather than of intervention effectiveness. Medline/OVID, Scopus, and EconLit will be searched, with no limitation on date. Relevant international policy websites will also be searched. The search is limited to papers in English. Citation tracing may identify further relevant papers. Abstracts and full texts will be screened using Rayyan. At least 10% will be double-screened, and the rest screened by one author, and included full texts will be screened until data saturation is reached. Study inclusion is based on consistency with the inclusion criteria, with some flexibility allowed due to the theoretical nature of this review. Data extracted from papers will be used to develop a diagrammatic map of pathways.

## Background and rationale

Access to adequate energy in the home is necessary for a healthy and well life, underpinning actions such as heating or cooling, cooking and storing food, lighting, washing and drying clothes, working from home, using medical equipment at home, and generally participating in society (Day, Walker and Simcock, 2016; Bouzarovski, 2018; Douglas *et al*., 2022). However, current consumption is unsustainable, with important implications for the mitigation of climate change (UNEP, 2017; Romanello *et al*., 2022). In the context of climate change, new energy vulnerabilities, such as the need to access additional energy to cool homes in a warmer climate, or to heat homes during extreme winter weather events, may increase energy consumption, reinforcing the effects of climate change and creating a positive loop between home energy use and climate change (Wilkinson *et al*., 2007). Decarbonising home energy is therefore an important step towards both reducing unsustainable energy use while maintaining quality of life (UNEP, 2017). Current projections for energy decarbonisation are not optimistic however, and the current overall rate of decarbonisation is much slower than required to meet key climate targets (Romanello *et al*., 2022).

Home energy decarbonisation can involve a variety of actions targeted at reducing energy consumption (for example, turning down lights, heating, or air conditioning), improving energy efficiency (for example, installing building insulation), and decarbonising energy at the source (for example, switching to renewable energy sources) (Wilkinson *et al*., 2009; Gibb, 2023). These actions can lead to benefits to health and wellbeing, for example, through improving thermal comfort or reducing indoor exposure to harmful pollutants (Wilkinson *et al*., 2007, 2009; UNEP, 2017). However, home energy decarbonisation may also have the potential to create new, or entrench existing, inequalities in health. There is evidence that those living in poorer quality housing may face higher fuel costs when warming or cooling their homes during extreme weather events than those living in better insulated buildings, producing an ‘energy inefficiency penalty’ for those less able to pay up-front costs to decarbonise their home energy (Age UK, 2023), with impacts to their health and wider health inequalities (Gibb, 2023).

### Research aims

The aims of this scoping review are to:

1. identify theorised causal pathways between decarbonisation of home energy and health and health inequalities, and
2. develop a putative causal evidence map of those pathways.

### Methodology

This protocol is for a review of theoretical pathways, rather than a review of intervention effectiveness, and the focus of the review is on exploring and understanding pathways and how they work together, rather than on gathering detailed information on their effectiveness. This review therefore aims to explore what happens between the exposure and outcome, rather than the impact of the exposure on a given outcome. Reviews of theories can be useful when drawing together literature across different disciplines, by identifying conceptual commonalities and providing a conceptual vocabulary for bridging disciplinary differences (Lorenc *et al*., 2012; Campbell *et al*., 2014). This can be useful to understanding how varied factors relate to each other, and for identifying causal pathways between those factors (Campbell *et al*., 2014), which can then be used as frameworks for further work. In the case of this review, taking a theory-led approach will be useful for identifying links between at least two distinct literatures, decarbonisation and energy, and health and health inequalities, and for the development of an evidence map of pathways to be used as a framework for later quantitative modelling.

### Inclusion criteria

Previous reviews of theories within health research have identified challenges in defining inclusion criteria that are both broad enough to include relevant literature but specific enough to be useful, as well as criteria that accurately capture ‘theory’ (Campbell et al., 2014). To counter this, studies have used both a priori inclusion criteria alongside more subjective methods of selecting papers, such as citation tracing, consultation with subject experts, and making informed subjective judgements (Lorenc et al., 2012; Bonell et al., 2013; Campbell et al., 2014). In their review of theories about the impact of the school environment on children’s health, Bonell et al. (2013) identified and included eight relevant reports that were not captured by their systematic search, noting that these were not previously identified due to date of publication, reference type, and lack of relevant key words. Similarly, in their review of pathways between crime, fear of crime, environment, and health, Lorenc et al. (2012) emphasised their use of ‘pearl growing’ methods such as citation tracing, following an initial search using specific inclusion criteria, to identify further relevant papers. In their paper outlining methodological challenges encountered whilst conducting a review of theoretical pathways between income and health, Campbell et al. (2014) note that key theoretical papers may not be specific to the areas of literature captured by inclusion criteria; this does not preclude the use of a priori inclusion criteria but requires an acknowledgement that flexibility in use of the criteria may be required. Drawing on these papers, the following section outlines a set of specific a priori inclusion criteria that will be used to search the literature to identify relevant papers. Citation tracing will also be conducted to identify other relevant literature that may not be captured by these terms.

The inclusion criteria for this review are outlined using a Population, Concept, Context (PCC) framework. There is no standard inclusion framework for reviews of theories (Campbell et al., 2014) and PCC is used here as it is often recommended for scoping and mapping reviews (Aromataris and Munn, 2020).

### Population

All populations within high-income countries are included, as defined by the World Bank in 2023/24 (World Bank, 2023). The focus on high-income countries is due to different energy needs and uses between different contexts (Day, Walker and Simcock, 2016; Pelz, Pachauri and Groh, 2018), plus the need to situate decarbonisation within the wider global challenge of reducing harmful emissions and unsustainable energy use equitably, and the responsibility of high-income countries to do this (Day, Walker and Simcock, 2016; Gillard, Snell and Bevan, 2017). Higher standards of living in high-income countries require more energy to sustain, thus more energy to decarbonise, and high-income countries have more access to more energy alongside more resources to distribute energy to the population (Day, Walker and Simcock, 2016).

### Concepts

#### Decarbonisation of home energy

Any intervention, policy or other action which achieves decarbonisation of household energy use and consumption, and which takes place in the home. This may include actions aimed at improving energy efficiency, reducing energy consumption (behaviours), or decarbonising energy sources, and is limited to energy use within the home. This excludes actions which may change the energy use of households in other areas e.g., transport or healthcare. Table 1 outlines some examples (Wilkinson et al., 2009; Gibb, 2023).

**Table 1.**
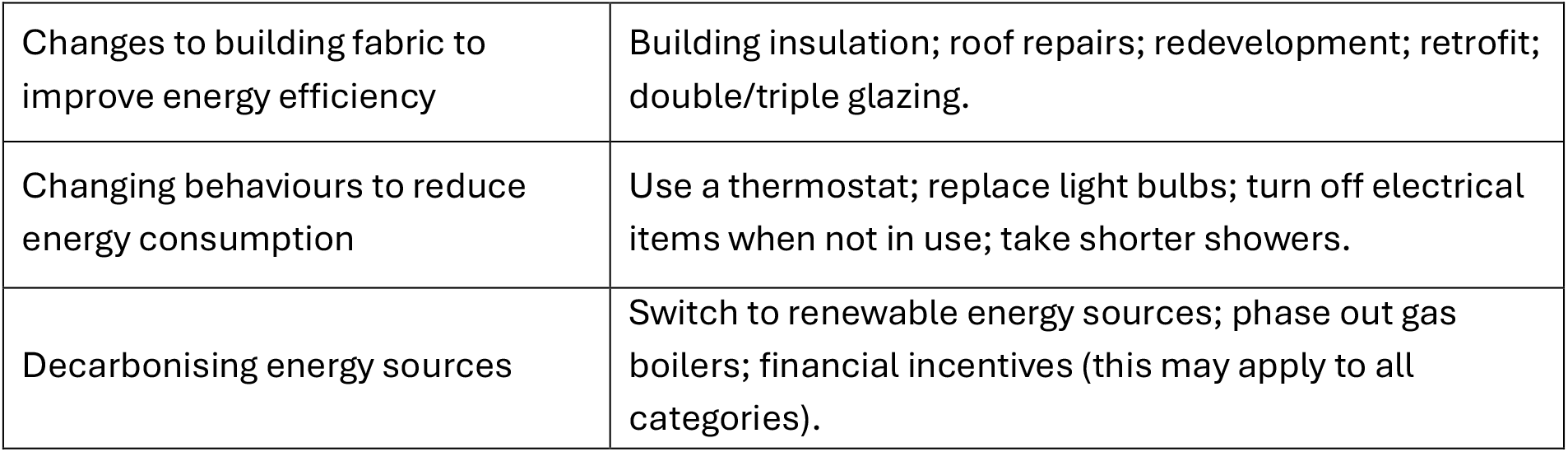
Example actions to decarbonise home energy consumption.

#### Health and health inequalities

Any physical or mental health or wellbeing outcome, and any measure of health inequality or inequity, for any physical or mental health or wellbeing outcome or measure. This includes any summary outcomes such as life expectancy, negative health outcomes such as mortality, and service use proxies such as hospitalisation and prescribing.

### Context: theorised pathway

Based on Campbell et al.’s (2014) inclusion criteria for their review of theoretical pathways between income and health, studies should incorporate a clear theory or hypothesis of *intervention* (in this case, decarbonisation of home energy) – *pathway* (mechanism of effect) – *outcome* (health and health inequalities).

### Study designs

All study designs are included, including quantitative, qualitative, theoretical, and reviews. Policy literature, i.e. reports produced by governmental organisations or non-governmental organisations, are included. Blogs, websites, opinion pieces, press releases, books, and newspaper articles will be excluded.

### Date

Any date.

### Language

Only papers written in English will be included.

### Search terms

Medline/OVID, SCOPUS and EconLit were searched on the 18^th^ July 2024 using the search terms listed in Table 2. Search terms for the context field (pathway) are adapted from the search strategy used by Campbell *et al*. (2014); concept (outcomes) search terms are adapted from those used by Boyd *et al*. (2022). Citation tracing will also be conducted where papers refer to additional papers which outline further pathways, or the same pathways in more depth.

**Table 2.** Search terms used to search Medline/OVID, SCOPUS and Econlit. Terms are for context: pathway, and concepts: decarbonisation of home energy, health, and health inequalities. Terms were searched for in abstract, keywords, and title.

|  |  |
| --- | --- |
| **Context:** theorised pathway | Theory or theories or pathway or model* or mechanism or mapping* or strategy or strategies or mitigation* |
| **Concept (1):** decarbonisation of home energy | (Decarboni* adj2 (home or domestic or hous* or heat* or cool* or energy or fuel)) or (energy adj2 (efficienc* or use* or renewable* or home)) or (retrofit* or retro fit*) or (insulation adj2 (cavity or wall or loft or internal)) or (green adj2 (hous* or energ* or redevelop* or renew*)) or (zero adj2 (net or emission*)) or (carbon adj2 (emission*)) or (low-carbon adj2 (home* or domestic or hous* or heat* or energy)) or (eco-friendly adj2 (home* or domestic or hous* or heat* or energy)) |
| **Concept (2):** health | sickness or health* or wellbeing or illness* or disabilit* or morbidit* or mortalit* or disease* or surviv* or hospital* or precrib* or “life expectancy” or death* or QALY or DALY |
| **Concept (2):** health inequalities | equit* or inequit* or inequalit* or disparit* or equality or socioeconomic or determinant* or outcome* or poverty or deprivation or low income or “socioeconomic disparities in health” or (social adj2 (class* or group*)) |

### Grey/policy literature search

Relevant international policy websites, listed below, were searched on the 18^th^ July 2024 using the terms ‘home energy’ and ‘health’. Only the first 20 results on each site will be included for screening.

- Energy and health, World Health Organisation: https://www.who.int/health-topics/energy-and-health/
- United Nations: https://www.un.org/en
- UN Environment Programme: https://www.unep.org/
- International Energy Agency: https://iea.org/
- C40 Cities: https://www.c40.org/
- Institute of Health Equity: https://instituteofhealthequity.org/
- UK Collaborating Centre for Housing Evidence: https://housingevidence.ac.uk/

### Data screening

All abstracts were uploaded to Zotero (Takats *et al*., 2024) on the 18^th^ July 2024. Abstracts were screened using Rayyan (Ouzzani *et al*., 2016). An initial sample of abstracts were screened by two reviewers to facilitate initial discussion of any challenges and/or differences encountered while screening. Following this, all abstracts were screened by one reviewer, with 15% screened by a second reviewer. Full texts will be screened by one reviewer, with at least 10% screened by a second reviewer. Full texts that meet the inclusion criteria will be reviewed until saturation of pathways is reached i.e., data saturation, which arises when no new pathways are named in the literature (Saunders *et al*., 2018).

Given that the focus of the review is to scope theoretical pathways rather than to systematically gather data on pathway effectiveness, reviewing until data saturation is reached avoids gathering redundant information on pathways that have been outlined or referred to repeatedly (Saunders *et al*., 2018). Screening will be double blinded on Rayyan to ensure independent decisions on paper inclusion/exclusion. Reviewers will meet regularly to discuss any differences and challenges.

### Data extraction

Data will be extracted according to the following categories: title, author(s), journal, year, study design, country of study, geographical setting/context, population, study term, intervention type and description, housing tenure, housing type, any other housing characteristics specified, name of pathway (if provided), summary of pathway, diagrammatic representation of pathway (if provided), health outcome(s) including inequalities, any other outcome(s), link to any other pathway (if provided). If provided, information on evidence to support the pathway including effect sizes will also be collected.

### Critical appraisal

Bonell *et al*. (2013) and Campbell *et al*. (2014) provide guidance on critical appraisal of theories. Given that the aim of this review is not to analyse or evaluate different pathways, but to scope and synthesise the literature around pathways, in-depth critical appraisal is outside the scope of this review and will not be undertaken. Included papers will be scored on their depth of theory, relevance to the research question, and originality (Bonell *et al*., 2013).

### Synthesis and outputs

Extracted data will be presented as a matrix using the categories listed above, which will be used to develop a causal map of pathways between intervention and outcome. The map will be used to inform a quantitative model testing one or several theoretical pathways to determine potential impacts of decarbonisation actions on health inequalities.

## Data Availability

All data used is from published journals and websites freely available online.

